# Investigating the oral and dental effects of nicotine pouches: findings from a patient and professional survey with in-person assessment

**DOI:** 10.64898/2026.09.29.26364219

**Authors:** CC Currie, JM Jackson, J Allison, P Basta, R Tapon, E Ash, N Lansdowne, N Atkinson, A Fucile, E Padgett, S Warner, S Miluna-Meldere, L Crombie, Z Toma, C Nile, N Jakubovics, R Kist, M Robinson, R Holliday

**Affiliations:** School of Dental Sciences, Newcastle University, Framlington Place, Newcastle Upon Tyne, NE2 4BW, UK; Newcastle Upon Tyne Hospitals NHS Foundation Trust, Newcastle Upon Tyne, UK; Population Health Sciences Institute, Faculty of Medical Sciences, Newcastle University, Newcastle upon Tyne, UK; Department of General Dentistry, Riga Stradins Univeristy, Dzirciema street 16, Riga, Latvia

## Abstract

**Background:** Nicotine pouches (NPs) are increasingly popular tobacco-free nicotine products, particularly among young people. Despite growing use, evidence regarding their oral health effects remains limited.

**Methods:** We employed a mixed-methods approach comprising: (1) a national cross-sectional survey of NP users, (2) a survey of UK dental professionals, and (3) in-person clinical assessments of NP users. Survey data explored patterns of NP use, oral and dental side effects, and perceptions of risk. Clinical assessments included comprehensive oral examinations, periodontal assessments, and histopathological investigation of lesions where indicated.

**Results:** A total of 3,659 NP users completed the public survey. Participants were predominantly male (89%) and aged 16-22 years (78%). Overall, 45% reported at least one oral or dental side effect. The most commonly reported effects were gingival recession (60%), burning sensations (41%), white patches (30%), and dry mouth (29%). Higher nicotine exposure was associated with increasing numbers of reported side effects. Among 126 dental professionals, oral lesions associated with NP use were commonly observed clinically, while only 17% reported seeing no oral or dental effects. Thirty-three NP users underwent in-person assessment. Ten lesions were biopsied; six demonstrated hyperkeratosis and four showed oral epithelial dysplasia (three mild, one moderate). All dysplastic lesions occurred at pouch placement sites. In two cases, clinical follow-up demonstrated partial or complete resolution of lesions following cessation of NP use. Periodontal assessments demonstrated generally low levels of periodontal disease and gingival recession across the cohort.

**Conclusions:** Oral and dental side effects associated with NP use were commonly reported by users and observed by dental professionals. Histopathological assessment identified biopsy-confirmed oral epithelial dysplasia in lesions occurring at pouch placement sites, representing, to our knowledge, the first such report associated with NP use. Evidence of lesion improvement following discontinuation of use suggests that at least some NP-associated oral changes may be reversible.

Although causality cannot be established, these findings highlight the need for vigilance among dental professionals and urgent longitudinal research to determine the incidence, progression, and reversibility of NP-associated oral lesions.

## Introduction

Nicotine pouches (NPs) are the latest addition to the rapidly evolving nicotine product landscape. Marketed as tobacco-free alternatives to traditional smokeless tobacco products, NPs are small, pre-portioned sachets containing nicotine, flavourings, sweeteners and plant-based fillers that are placed between the lip and gingiva, allowing nicotine absorption through the oral mucosa(Jackson et al., 2023).

Their discreet use, wide range of flavours and relatively unrestricted availability have contributed to substantial growth in popularity across many countries, with epidemiological data highlighting increased usage amongst young populations (Mahmoodianfard et al., 2026). In the United Kingdom (UK), overall rates of NP use have increased from 0.1% in 2020 to 1.0% in 2025, with a particularly high prevenance among young people (aged 16-24 years) (Tattan-Birch et al., 2026). This trend is concerning for public health professionals and regulators; not only because of the systemic effects of NP use, such as cardiovascular risks (Dennison Himmelfarb et al., 2025), but because of the addictive qualities of nicotine and the potential targeting of young people through NP’s flavour profiles, packaging, and marketing strategies. In response, France has already implemented a total ban of NP products in April 2026 (Casassus, 2025).

Compared with the growing body of literature examining the behavioural and toxicological aspects of NP use (Al-Otaibi and Althobiani, 2025, Tattan-Birch et al., 2026, Rungraungrayabkul et al., 2024), considerably less is known about their effects on the oral cavity. Early studies have reported localised mucosal changes at pouch placement sites, including white lesions, mucosal irritation, epithelial thickening and gingival recession (Masoumeh et al., 2026, Alkhatib, 2025, Dowd et al., 2024) (REF). However, the current evidence base remains limited; largely comprising cross-sectional observations and case reports, with no published biopsy- confirmed cases of oral epithelial dysplasia associated with NP use. Consequently, the pathological significance of many NP-associated oral lesions remains uncertain.

Given the increasing prevalence of NP use and the paucity of data regarding their oral health effects, further clinical investigation is required. Therefore, the aim of this study was to explore the oral and dental effects of NP use and understand the best methodological approaches to study these effects.

## Methods

This study received approval from the UK Health Research Authority and NHS Research Ethics Committee.

### Public survey

A cross-sectional survey was conducted online using JISC Online Surveys. The survey was promoted via several routes including: local posters mainly around Newcastle Upon Tyne, media stories (Vlope, 2026), social media posts, distribution by relevant organisations (e.g. NuTH (National Health Service, 2026)), and paid advertising on Facebook and Instagram that was utilised via Meta Platforms Inc..

Inclusion criteria were individuals aged 16 years and above who self-reported the use of oral NP’s . Exclusion criteria were participants who were unable to read or speak English or who were unable to provide valid consent. Structured questions were asked which covered participant demographics, nicotine use including dosage of products, motivations for use, and self-reported oral health side effects (A copy of the survey instrument can be viewed in Supplementary Material). The survey also included space for open-ended responses. Participants had the option to upload a photograph of any lesions through their own smart phone, or an intra-oral photograph device (DentalMonitoring Scanbox Pro and Software) to aid in taking the photograph. All survey participants were entered into a prize draw to win £100.

### Dental professional survey

A cross-sectional survey of dental professionals was also conducted, where questions focused on participants’ knowledge on smoking, NPs, and patient experiences of NP use. This was conducted via JISC Online Surveys. The survey was promoted via several routes including: in academic journals (Br Dent J, 2026, Br Dent J Team, 2026), posting on relevant social media groups and by paid advertising by Meta Platforms Inc, which supported adverts on Facebook and Instagram.

Inclusion criteria included those registered with the General Dental Council as a dentist, dental therapist, dental hygienist, dental nurse, orthodontic therapist, or clinical dental technician, and who primarily practised dentistry in the United Kingdom. The exclusion criterion was membership of the research team. All survey participants were entered into a prize draw to win £100.

### In-person assessments

Public survey participants were asked whether, if selected, they would attend an in-person assessment at the Newcastle Dental Clinical Research Facility. A comprehensive oral and dental examination was completed, biological samples collected (saliva, dental plaque, dental calculus, gingival crevicular fluid, oral swabs and brush biopsies) and where indicated, participants were offered an incisional biopsy of any lesions noted. These were carried out under local anaesthetic by two members of the research team (JJ, RH). Biopsy specimens were examined by two members of the research team (CC, MR) using standard H&E stains, with additional diastase resistant PAS staining as indicated. Presence of oral epithelial dysplasia was graded using the World Health Organization (WHO) classification of mild, moderate or severe(Odell et al., 2021). Participants were offered a £50 voucher for completion of in-person assessments.

### Analysis

#### Public and dental professional survey

Data from both surveys were downloaded into Microsoft Excel for data cleaning, followed by analysis in STATA version 15 (StataCorp LLC, College Station, TX, USA). Free text responses were reviewed, and responses were re-categorised as required by one member of the research team (CC).

Self-reported dose data were converted into three variables, one for responses in milligrams, one for responses in “dot”, and one with all dosage data converted into milligrams (further details are in the Supplementary Appendix). Data were analysed using descriptive statistics. Consideration of a dose-effect relationship was explored using Mann-Whitney, Spearman rank, linear and logistic regression modelling.

#### In person assessment

Participant characteristics were summarized descriptively using SPSS (Version 31.0.1.0 [49]). Nonparametric statistical hypothesis tests (Wilcoxon rank-sum test with exact *P* values) compared continuous periodontal parameters between pouch and non-pouch sites.

## Results

### Public survey

A total of 3,663 responses were received for the public survey. Following data cleaning, four responses were excluded due to answers consistent with those generated by AI, leaving a total of 3,659 responses for analysis.

Participant demographics and NP use are summarised in Table 1. The majority of participants were current NP users (92%), while 8% reported former NP use. Of these, most participants were male (89%), White British (69%) and aged 16 - 22 years (78%). Among participants with a postcode that could be linked to deprivation data (n=2,648), representation was observed across all Index of Multiple Deprivation (IMD) deciles, although a slightly greater proportion resided in less deprived areas (deciles 7-10). Half of respondents reported being in full-time education and 39% were employed.

**Table 1:** Characteristics of participants from public survey.

| **Characteristic** | **n (%)** |
| --- | --- |
| **Age group (years)** |  |
| 16-18 | 1,319 (36) |
| 19-22 | 1,538 (42) |
| 23-25 | 347 (9) |
| 26-30 | 259 (7) |
| 31-40 | 157 (4) |
| 41-50 | 31 (1) |
| 51-60 | 5 (<1) |
| 61-70 | 2 (<1) |
| 71-80 | 1 (<1) |
| **Gender** |  |
| Male | 2,993 (89) |
| Female | 342 (10) |
| Non-binary | 16 (0.5) |
| Prefer not to say | 14 (0.5) |
| Trans female | 1 (<0.5) |
| **Ethnicity\*** |  |
| White - British | 2,531 (69) |
| Any other White background | 246 (7) |
| White - Irish | 122 (3) |
| Mixed – White and Asian | 101 (2) |
| Asian/Asian British - Indian | 77 (2) |
| Missing | 292 (8) |
| **Nicotine pouch status** |  |
| Current NP user | 3,367 (92) |
| Former NP user | 292 (8) |
| **Other nicotine products used** |  |
| E-cigarette user | 1,654 (45) |
| Cigarette smoker | 1,443 (39) |
| Other nicotine products | 107 (3) |
| **Employment status** |  |
| Full-time education | 1,686 (50) |
| Employed | 1,308 (39) |
| Unemployed | 261 (8) |
| Self-employed | 107 (3) |
| Retired | 5 (0.2) |
| **IMD Decile\*\*** |  |
| 1 | 130 (5) |
| 2 | 168 (6) |
| 3 | 211 (8) |
| 4 | 271 (10) |
| 5 | 247 (9) |
| 6 | 297 (11) |
| 7 | 359 (13) |
| 8 | 326 (12) |
| 9 | 347 (13) |
| 10 | 364 (14) |
| **Duration of NP use** |  |
| <1 month | 35 (1) |
| 1-6 months | 407 (14) |
| 6 months-1 year | 555 (19) |
| 1-2 years | 1,152 (39) |
| 2-5 years | 747 (25) |
| >5 years | 65 (2) |
| **Pouches used per day** |  |
| 1 | 155 (5) |
| 2-5 | 1,556 (53) |
| 6-10 | 998 (34) |
| 11-20 | 241 (8) |
| >20 | 11 (0.4) |
| **No. of hours used per day** |  |
| <1 | 610 (21) |
| 1-2 | 1,017 (35) |
| 3-4 | 701 (24) |
| 5-6 | 273 (9) |
| 7-8 | 155 (5) |
| 9-10 | 73 (2) |
| 11-12 | 54 (2) |
| 13-14 | 33 (1) |
| 15-16 | 26 (1) |
| >16 | 19 (<1) |

\*A wide range of ethnicities were included therefore only the most frequently reported groups are shown.
\*\*IMD is the official measure of deprivation in the UK and considers deprivation being related to more than just poverty. IMD combines seven different domains: income; employment; health deprivation and disability; education, skills and training; crime; barriers to housing and services; living environment. There are 32,844 lower layer super output areas (LSOAs) in England, with each being assigned a ranked IMD score, with 1 being the most deprived area, and 32,844 being the least deprived. For the purposes of this study IMD was considered in deciles, decile 1 is the most deprived and or decile 10 the least deprived. IMD data were available only for participants providing a postcode that could be matched to an IMD decile.

Concurrent use of other tobacco and nicotine products was common, with 45% reporting e-cigarette use, and 39% cigarette smoking. Only 39% reported exclusive NP use, whereas 21% used NPs in combination with both cigarettes and e-cigarettes. NP use occurred throughout the day, with the most commonly reported time periods being the afternoon (63%), early evening (63%), and lunchtime (61%). Patterns of use across the day indicated that many participants used NPs repeatedly. More than one-quarter (27%) reported use at all six time points (morning, mid-morning, lunchtime, afternoon, early evening, and night-time). Participants most commonly reported buying NPs at shops (71.5%) or online (26.8%), other less common places of purchase included: friends and family and social media.

Participants were asked about their opinions of NPs in terms of their knowledge of contents, risks and benefits; the majority revealed that they needed more knowledge about these products (Appendix Figure 1).

The median dose of NP used was 17mg or 4 dots. Higher nicotine dose was associated with the presence of white patches (z = −5.55, p < 0.001) and gingival recession (z = −3.44, p = 0.001). No significant associations were observed between dose and other oral/dental side effects (all p > 0.05). Using logistic regression, higher nicotine exposure was statistically associated with oral white patches and gingival recession, but the observed effect sizes were very small, suggesting limited clinical significance. In adjusted analyses, cumulative nicotine dose remained independently associated with gingival recession, whereas no significant association was observed for oral white patches (Further details in Supplementary Appendix).

Overall, 1,635 participants (45%) reported experiencing at least one oral or dental side effect associated with NP use, summarised in Table 2. The most common location reported for all side effects was upper labial mucosa, consistent with the most common reported site of NP use (labial mucosa – 65% and buccal mucosa – 60%). Respondents reporting increasing numbers of oral/dental side effects were positively correlated with NP dose. Weak but statistically significant associations were observed between increasing numbers of side effects and both NP dose (Spearman’s ρ = 0.052, p = 0.005) and estimated daily nicotine dose (Spearman’s ρ = 0.114, p < 0.001). A similar association was observed for number of pouches used per day (Spearman’s ρ = 0.112, p < 0.001) and cumulative dose (Spearman’s ρ = 0.099, p < 0.001). In linear regression analyses, greater nicotine exposure was associated with a higher number of self-reported side effects. These associations remained significant after adjustment for age, sex, cigarette smoking, and e-cigarette use, although effect sizes were very small and of uncertain clinical relevance (Supplementary Appendix).

**Table 2:** Reported oral and dental side effects of NP use in public survey. Data for severity and onset are not available for the four additional side effects identified from free text responses.

| Side Effect | N (%) | Mean Severity | Onset | Median Dose |
| --- | --- | --- | --- | --- |
| Gingival recession | 988 (60%) | 5 (SD 2.6) | After use (54%) | 26mg |
| Burning sensation | 662 (41%) | 3 (SD 1.9) | During use (76%) | 17mg |
| White patches | 494 (30%) | 4 (SD 2.4) | After use (45%) | 20mg |
| Dry mouth | 470 (29%) | 4 (SD 2.1) | Immediately after use (37%) | 17mg |
| Ulcers | 302 (19%) | 5 (SD 2.4) | After use (65%) | 16mg |
| Sore throat | 229 (14%) | 4.5 (SD 2.1) | After use (42%) | 17mg |
| Sore mouth | 220 (13%) | 4 (SD 2.1) | After use (51%) | 17mg |
| Gingival sloughing | 74 (5%) |  |  |  |
| Bleeding gums | 31 (2%) |  |  |  |
| Halitosis | 14 (1%) |  |  |  |
| Tooth sensitivity | 13 (1%) |  |  |  |

Just over half (52%) of respondents reported systemic side effects, the most common being headaches (45%), vomiting (38%), difficulty sleeping (32%), anxiety (29%) and chest tightness (19%). The majority had not seen a healthcare professional about these. Of those who had consulted a healthcare provider (3%) the most common presentation was to a GP (63%). Others included dentists, emergency departments, cardiologists, and pharmacists.

Mint was the predominant flavour choice, reported by 40% of participants. A chi-square test demonstrated a significant association between flavour category (mint versus non-mint, details in supplemental appendix) and the presence of oral or dental side effects, χ²(1)=5.91, p=0.015. However, when flavour was examined within a multivariable logistic regression model controlling for dose per day and duration of use, mint flavour was not independently associated with oral side effects (OR=1.01, 95% CI 0.87-1.17, p=0.858).

### Dental professional survey

A total of 126 dental professionals completed the survey; general dental practitioners (41%), dental nurses (20%), dental hygienists (14%), specialist/specialty trainees (10%), dental therapists (10%), and other hospital grade dentists (5%).

Over one-half (58%) reported routinely asking patients whether they use NP’s, and just under one-half (45%) included this within the medical history form. Participants most commonly reported seeing either 1-5 NP patients (33%) or more than 30 patients (30%) in the previous year. The majority (42%) reported that patients did not complain of any side effects associated with NP use. However, when asked about changes noted during clinical examination, only 17% reported seeing no oral or dental effects from NP use. Oral and dental side effects noted on clinical examination are shown in Appendix Figure 2. Of note 3% (n=4) reported patients having biopsies of these lesions showing mild or low-grade epithelial dysplasia.

The majority of participants believed that NPs are either less harmful (68%) or about the same (24%) as tobacco cigarettes, whereas the majority believed that they are more harmful (38%) or about the same (33%) as e-cigarettes. Interestingly, 41% believed that NPs are carcinogenic, while 48% were unsure. Further, the majority of participants reported little of no confidence in understanding and discussing the effects of NPs with patients (Appendix Figure 3).

Participants provided free text responses relating to advice / management strategy for discussing NP with patients, carcinogenic concerns, and additional concerns of NP usage (Examples of these comments can be viewed in Appendix Tables 5-7).

### In-person assessment

In total, 783 survey respondents noted interest in taking part in the in-person assessment. After assessing eligibility (based on location), 277 were invited, which resulted in 33 attending the Newcastle Dental Clinical Research Facility between 8^th^May 2026 and 11^th^ September 2026. The participant demographics are shown in Appendix Table 8, which were similar to the survey data (Table 1); predominately young males, with a high concurrent use of nicotine and other tobacco products.

Periodontal indices generally showed low levels of periodontal disease/conditions. Comparison between pouch and non-pouch sites showed statistically significant differences for Periodontal Pocket Depth (PPD), percentage of sites with Clinical Attachment Level (CAL) ≥3mm, percentage of sites with Recession (REC) ≥2mm, (Bleeding on Probing (BOP), and Plaque Index (PI), with pouch sites always having lower scores (see Appendix Table 9).

### Histopathological findings

Ten participants presented with soft tissue lesions, which were judged to warrant formal biopsy (Figure 1). Following histological examination six participants were found to have hyperkeratosis only. In all cases, the specimens the squamous epithelium showed hyperparakeratosis, which was often markedly thickened (Figure 3). The epithelium was either of normal thickness for the site or hyperplastic. There was largely minimal to no inflammatory response, however one case showed background focal lichenoid inflammation. None of the specimens demonstrated presence of fungal hyphae using diastase resistant PAS staining.

Four cases showed evidence of epithelial dysplasia, three graded as mild and one as moderate (Figure 2).(Odell et al., 2021) In all cases, the squamous epithelium showed hyperparakeratosis and was either normal thickness for the site or was atrophic. Additionally, all cases had basal cell hyperplasia with a marked increase in the number of para-basal and supra-basal mitotic figures. In three cases, there was patchy background lichenoid inflammation, and none of the specimens demonstrated presence of fungal hyphae using diastase resistant PAS staining.

## Discussion

This study provides new evidence regarding the oral health effects of NP use and, to our knowledge, reports the first biopsy-confirmed cases of oral epithelial dysplasia identified in association with NP-related oral lesions. Almost half of survey respondents reported at least one oral or dental side effect, with gingival recession, burning sensations and white patches being most common. These findings were supported by reports from dental professionals and detailed clinical assessments.

Histopathological examination identified epithelial dysplasia in four lesions, including one moderate-grade lesion. Although causality cannot be established from this observational study, several findings support this, including lesion occurrence at pouch placement sites, improvement following cessation or relocation of pouch use, and evidence of dose-response relationships. Collectively, these findings suggest that oral lesions associated with NP use warrant clinical attention and further investigation.

The mechanisms responsible for these lesions remain unclear. NP products vary considerably in nicotine concentration, flavourings, pH modifiers and other constituents, any of which may contribute to local tissue injury. We observed evidence of a dose-effect relationship for some self-reported outcomes such as white patches and gingival recession. Our in-person assessment observed minimal levels of oral diseases/conditions. We found conflicting findings to a recent study on firefighters using NPs; our study found pouch sites had significantly lower recession and clinical attachment loss whilst the other study found significantly higher levels of clinical attachment loss in NP users verses non-users (Chaffee et al., 2026). The difference could perhaps be explained by different methodologies (comparison at site or participant level) and the different demographics with our participants being on average approximately a decade younger.

We also found discrepancies between public and professional reports of oral lesions. White patches were reported substantially more frequently by dental professionals than NP users, suggesting that self-reported surveys may underestimate lesion prevalence. This is clinically important because white lesions were the most common presentation among biopsied sites and were present in all dysplasia cases. Users may therefore be unaware of potentially significant oral changes, highlighting the importance of regular dental attendance and careful soft tissue examination. Notably, in several cases where lesions appeared clinically as likely hyperkeratosis, histopathological examination showed the presence of epithelial dysplasia, suggesting that a low threshold for histopathological assessment of persistent pouch-site lesions may be sensible.

Comparisons with e-cigarettes are particularly relevant. Despite almost two decades of widespread e-cigarette use, evidence of detrimental oral effects remains minimal (Tattar et al., 2025, Holliday et al., 2021) particularly of clinically significant oral mucosal pathology. By comparison, NPs are a relatively recent addition to the nicotine marketplace, yet we identified epithelial dysplasia within lesions arising at pouch placement sites. While causality cannot be inferred, the prolonged and repeated direct contact between NPs and oral tissues represents a fundamentally different exposure pattern to vaping and may explain the site-specific pathological changes observed.

Our findings extend a small but growing literature describing oral effects associated with NP use. Previous studies have reported mucosal irritation, white lesions and gingival recession, but evidence has largely been limited to case reports, cross-sectional observations, and self-reported symptoms. However, no previous study has combined large-scale surveys with comprehensive clinical assessment and histopathological investigation. Most importantly, to our knowledge, oral epithelial dysplasia has not previously been reported in association with NP use. These findings therefore move the evidence base beyond reversible mucosal irritation and show that, in some users, prolonged exposure may be associated with clinically significant pathological changes.

Several limitations should be considered. The public survey relied on self-reported data and is therefore susceptible to recall, reporting, and selection biases. The cross-sectional design prevents causal inference and residual confounding remains possible. In particular, many participants reported current or former cigarette smoking and e-cigarette use, both of which may have influenced observed findings. Several participants with dysplastic lesions had current or previous smoking histories, although one dysplasia case occurred in a never-smoker, suggesting established risk factors alone are unlikely to fully explain the findings. The in-person sample was relatively small, and comparisons between pouch and non-pouch sites should be interpreted cautiously because anatomical differences between sites may influence periodontal measurements. Nevertheless, the study benefits from a large sample of NP users, triangulation of findings across users and dental professionals, detailed clinical examinations and histopathological confirmation of lesions.

These findings have important implications for clinical practice and public health. NP use was concentrated among young people, reflecting growing uptake in younger populations. Both users and dental professionals reported uncertainty regarding the oral health effects of these products and expressed a desire for further information. Clinicians should routinely enquire about NP use, undertake careful soft tissue examination, and maintain a low threshold for biopsy of lesions occurring at pouch placement sites. Further longitudinal studies are urgently required to determine the incidence, progression, and reversibility of NP-associated lesions, identify causative mechanisms, and inform evidence-based clinical and regulatory guidance.

## Supporting information

Dental Professional Survey

Public Survey

## Data Availability

All data produced in the present study are available upon reasonable request to the authors

## Acknowledgements

This independent research was funded by the Crispian Scully Research Award from the Oral and Dental Research Trust (ODRT) and the British Society for Oral and Dental Research (BSODR) and was delivered through the National Institute for Health and Care Research (NIHR) Newcastle Dental Clinical Research Facility (CRF) with additional support from the NIHR Biomedical Research Centre: Newcastle (BRC) awarded to the Newcastle upon Tyne Hospitals NHS Foundation Trust, Newcastle University and Cumbria, Northumberland, Tyne and Wear Foundation Trust. The views expressed are those of the author(s) and not necessarily those of the ODRT/BSODR, the NIHR or the Department of Health and Social Care.

AI Disclosure: Microsoft Copilot (Microsoft Corporation, Redmond, WA, USA) was used to assist with manuscript editing and organization and to support the standardization of nicotine pouch dose data according to predefined investigator- derived conversion rules. All AI-generated outputs were reviewed and verified by the authors, who accept full responsibility for the final manuscript and analyses.

## Author Contributions

C.C. Currie and J.M. Jackson contributed equally to this work and contributed to conceptualization, methodology, investigation, resources, data curation, formal analysis, visualization, funding acquisition, and writing of the original draft. J. Allison, P. Basta, and R. Tapon contributed to investigation, methodology, resources, and writing review and editing. N. Lansdowne, N. Atkinson, A. Fucile, and E. Padgett contributed to investigation, resources, and writing review and editing. S. Warner contributed to investigation, project administration, resources, and writing review and editing. S.M. Meldere contributed to methodology, resources, and writing review and editing. L. Crombie and Z. Toma contributed to investigation, methodology, resources, and writing review and editing. C. Nile contributed to investigation, methodology, funding acquisition, and writing review and editing. N. Jakubovics and R. Kist contributed to methodology, funding acquisition, and writing review and editing. M. Robinson contributed to investigation and writing review and editing. E. Ash contributed to writing of the original draft and writing review and editing. R. Holliday contributed to conceptualization, methodology, formal analysis, data curation, supervision, project administration, funding acquisition, writing of the original draft, and writing review and editing.

All authors critically reviewed the manuscript, approved the final version, and agree to be accountable for all aspects of the work.

---

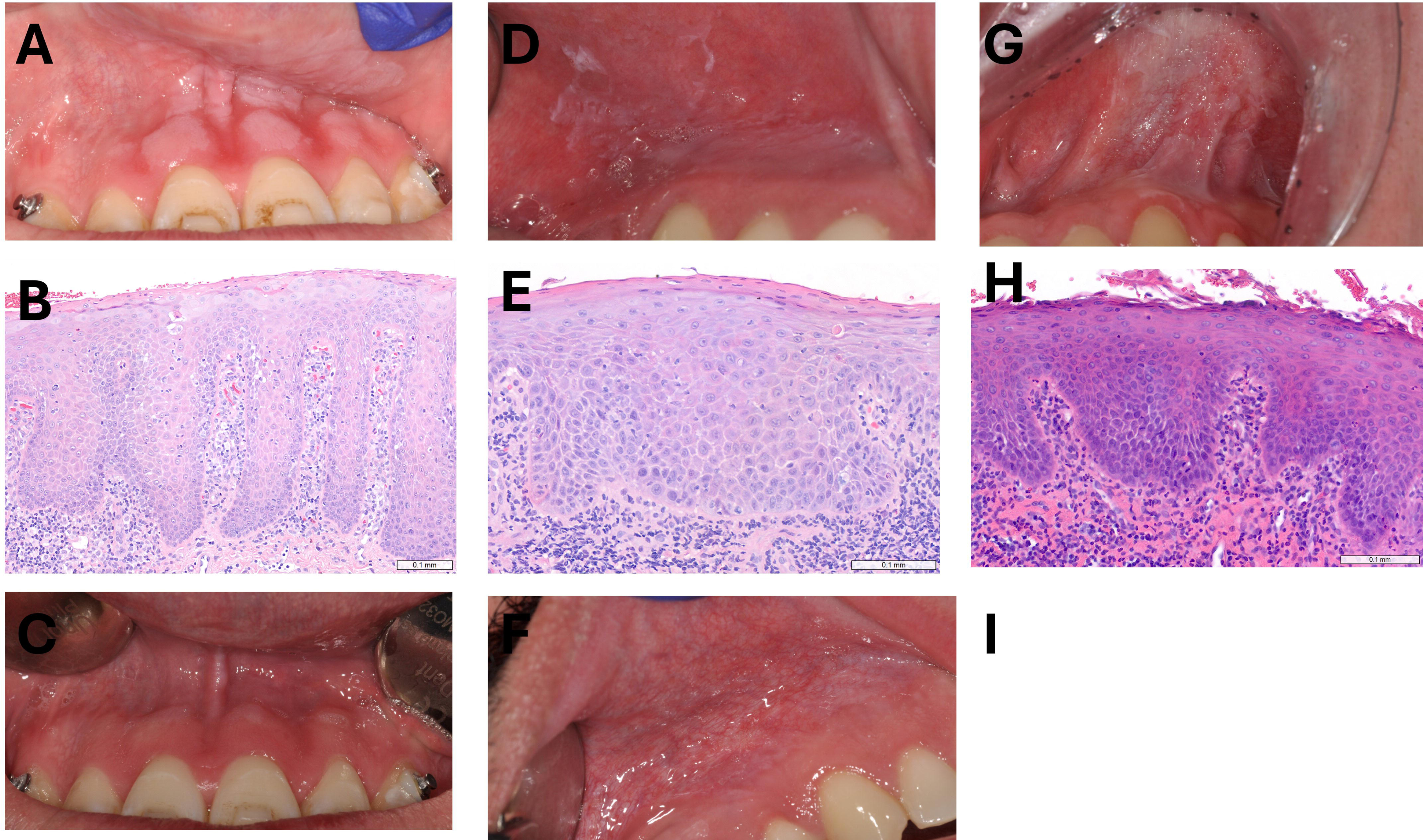

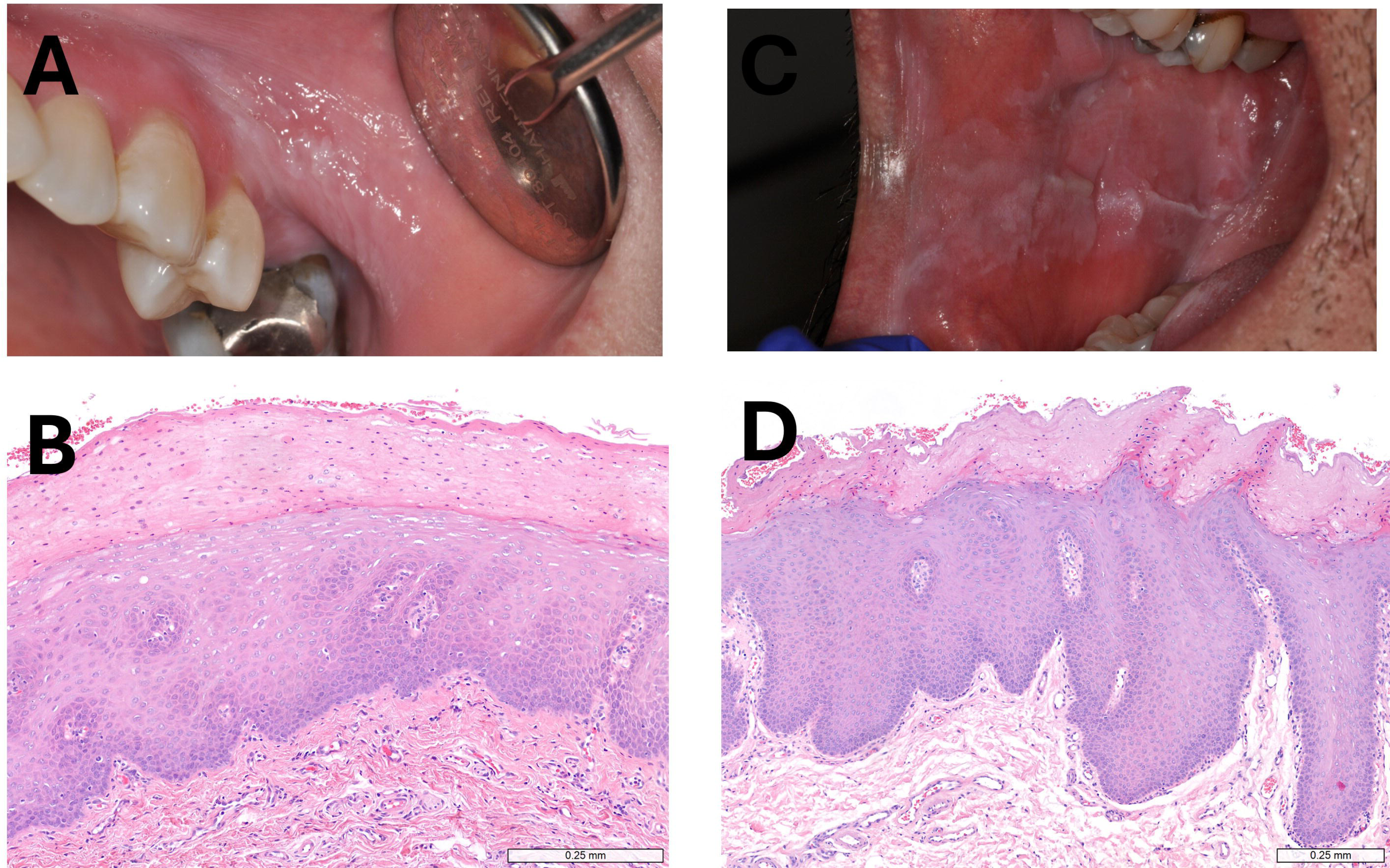

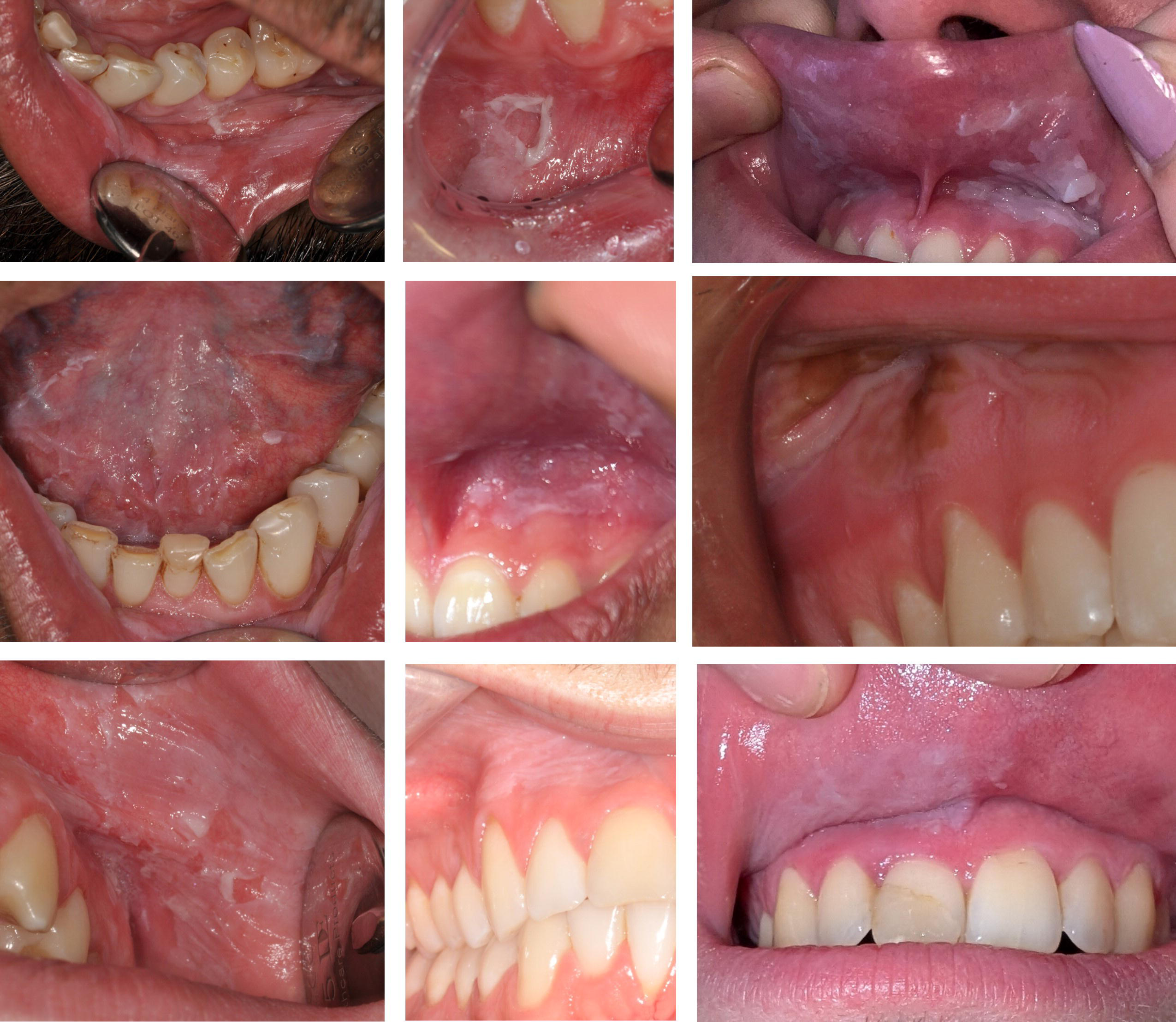

