## Supplemental Appendix for "Investigating the oral and dental effects of nicotine pouches: findings from a patient and professional survey with in-person assessment"

### 1. Additional Methods

**Generation of a standardised mg dose**

Where the NP dose was reported using qualitative descriptors (e.g. dot rating, "strong", or "max"), equivalent nicotine doses (mg) were assigned using predefined brand-specific conversion rules as shown below. Microsoft Copilot was used to assist in applying these conversion rules to the dataset, and all assignments were reviewed and verified by the investigators.

Dot numbers were changed to single values (e.g. 1 dot = 1, 2 dots = 2). Qualitative units were changed to single values (“strong” = 4, “extra strong/X-strong” = 5, “max” = 6). All dot values were then converted to mg using the brand given as follows:

- Velo:
  - 1 dot = 4mg
  - 2 dots = 6mg
  - 3 dots = 8mg
  - 4 dots = 10-11mg
  - 5 dots = 14mg
  - 6 dots = 17mg
- Nordic Spirit:
  - Regular = 6mg
  - Strong = 9mg
  - X-Strong = 11mg
  - Max = 17mg
- ZYN:
  - Low=1.5-3mg
  - Medium = 6mg
  - Strong = 9mg
  - X-Strong = 11mg
  - Max/XX-Strong = 13.5-16.5mg
- Pablo:
  - Silver = 10mg
  - Gold = 17mg
  - Original Ice cold = 20-24mg
  - Exclusive = 30mg

Where more than one dose was given as a free text response the first dose given was used (e.g. “4 dots, previously 6 dots” - 4 dots used for analysis). Once all doses were converted into a standardised mg dose additional variables were created in STATA to consider the dose effect. Estimated daily dose was calculated as the mg dose used multiplied by the median of the range given for number of times NPs were used per day (e.g. 2-5 pouches per day = 4, 6-10 pouches per day = 8). Cumulative dose was calculated as estimated daily dose multiplied by the median of the range of duration of use (e.g. 1-6 months = 3.5 months, 11-20 months = 15.5 months).

### 2. Additional Results

#### Public Survey

**Opinions on NPs**

When asked about their opinions of NPs the majority of respondents agreed that they understood what NPs contain and what the risks and benefits of their use are. However, the majority also agreed that they would like to know more about these, as highlighted in Appendix Figure 1 below. The majority of users reported learning about NPs online (67%), on social media (43%), from friends or family (33%) or at school (8%).

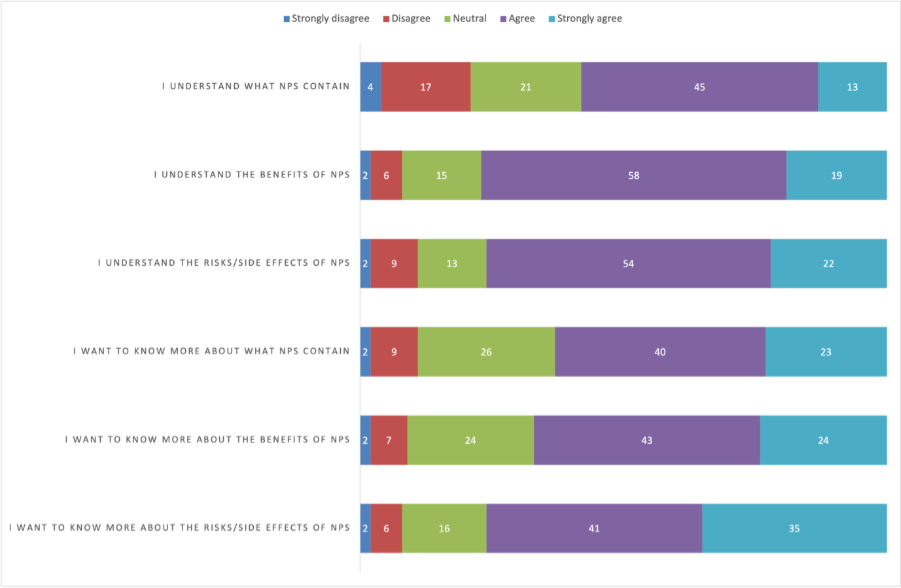

*Appendix Figure 1: User opinions on nicotine pouches.*

**Regression analysis for dose-effect**

Using logistic regression, higher daily nicotine dose was significantly associated with the presence of oral white patches (OR 1.001 per unit increase, 95% CI 1.0002-1.0012, p = 0.005) and gingival recession (OR 1.001 per unit increase, 95% CI 1.0004-1.0014, p < 0.001). When cumulative dose (estimated daily dose × duration of use) was examined, the association remained significant for gingival recession (OR 1.000017, 95% CI 1.000004-1.000029, p = 0.009) but not for white patches (OR = 1.000008, 95% CI 0.9999997-1.0000168, p = 0.058).

In multivariable logistic regression analyses (Appendix Table 1) adjusted for age group, gender, cigarette smoking, and e-cigarette use, cumulative dose remained independently associated with gingival recession (aOR 1.000025, 95% CI 1.000009-1.000041; p=0.002). No significant associations were observed for concurrent cigarette or e-cigarette in either model.

*Appendix Table 1: Multivariate logistic regression model for cumulative NP exposure and presence of white patches and gingival recession.*

| **Variable** | **White patch aOR (95% CI)** | **p-value** | **Gingival recession aOR (95% CI)** | **p-value** |
| --- | --- | --- | --- | --- |
| **Cumulative** | **1.000012** (0.999998-1.000012) | **0.094** | **1.000025** (1.000009-1.000041) | **0.002** |
| **Age group (** |  |  |  |  |
| 26-40 y | 0.57 (0.38-0.85) | 0.006 | 0.73 (0.56-0.97) | 0.028 |
| ≥41 y | 0.76 (0.22-2.57) | 0.656 | 0.10 (0.01-0.70) | 0.021 |
| **Gender (** |  |  |  |  |
| Woman | 0.95 (0.62-1.46) | 0.821 | 1.30 (0.95-1.77) | 0.099 |
| Non-binary | 1.40 (0.16-12.37) | 0.763 | 0.69 (0.08-6.15) | 0.738 |
| Prefer not to say | 1.08 (0.12-9.52) | 0.943 | 1.21 (0.22-6.73) | 0.831 |
| **Current cigarette use** | 1.22 (0.85-1.74) | 0.280 | 1.22 (0.92-1.61) | 0.161 |
| **Current e-cigarette use** | 1.22 (0.86-1.73) | 0.275 | 1.03 (0.78-1.35) | 0.853 |

**Regression analysis for self-reported side effects**

In linear regression analyses, cumulative nicotine pouch dose was positively associated with increasing numbers of self-reported side effects (Appendix Table 2). In the unadjusted model, cumulative dose was associated with increasing numbers of effects (β=0.0000057, 95% CI 0.0000007-0.0000106; p=0.025). After adjustment for age group, gender, cigarette smoking, and e-cigarette use the association remained significant (β=0.0000068, 95% CI 0.0000006-0.0000129; p=0.032). Similar findings were observed when dose was analysed as daily nicotine dose rather than cumulative dose (β = 0.00026, 95% CI 0.000001-0.00051; p = 0.049) (Appendix Table 3).

*Appendix Table 2: Adjusted linear regression model for number of self-reported oral and dental side effects related to cumulative exposure. Model statistics: R² = 0.025; N = 1,870.*

| **Variable** | **β coefficient (95% CI)** | **p-value** |
| --- | --- | --- |
| **Cumulative exposure*** | **0.0000068 (0.0000006 to 0.0000129)** | **0.032** |
| **Age group (ref: 18-25 years)** |  |  |
| 26-40 years | -0.182 (-0.335 to -0.030) | 0.019 |
| ≥41 years | -0.453 (-0.944 to 0.037) | 0.070 |
| **Gender (ref: man)** |  |  |
| Woman | 0.273 (0.090 to 0.455) | 0.003 |
| Non-binary | -0.392 (-1.443 to 0.658) | 0.464 |
| Prefer not to say | 0.262 (-0.786 to 1.310) | 0.624 |
| **Current cigarette use** | 0.059 (-0.101 to 0.220) | 0.469 |
| **Current e-cigarette use** | 0.124 (-0.033 to 0.282) | 0.122 |

*Appendix Table 3: Adjusted linear regression model for number of self-reported oral and dental side effects related to daily nicotine dose. Model statistics: R^2^ 0.027, n=1870.*

| **Variable** | **β coefficient (95% CI)** | **p-value** |
| --- | --- | --- |
| **Dose per day** | 0.00026 (0.000001 to 0.00051) | 0.049 |
| **Duration of use (months)** | 0.00406 (0.00036 to 0.00775) | 0.031 |
| **Age group (ref: 18-25 years)** |  |  |
| 26-40 years | -0.197 (-0.351 to -0.044) | 0.012 |
| ≥41 years | -0.458 (-0.948 to 0.032) | 0.067 |
| **Gender (ref: man)** |  |  |
| Woman | 0.295 (0.112 to 0.479) | 0.002 |
| Non-binary | -0.411 (-1.460 to 0.638) | 0.442 |
| Prefer not to say | 0.283 (-0.764 to 1.330) | 0.596 |
| **Current cigarette use** | 0.063 (-0.098 to 0.223) | 0.443 |
| **Current e-cigarette use** | 0.126 (-0.031 to 0.283) | 0.116 |

Users of mint-flavoured NP’s reported slightly more oral and dental side effects than users of non-mint flavours (mean 1.02 vs 0.92; t = 2.21, p = 0.027). In a multivariable linear regression model adjusting for nicotine dose and duration of use (Appendix Table 3), mint flavour remained independently associated with more self-reported side effects (β = 0.092, 95% CI 0.004 to 0.181, p = 0.041), although the overall effect size was small (R² = 0.006).

*Appendix Table 4: Oral or dental side effect of NP flavour*

| **Flavour** | **No oral or dental side effect** | **Oral or dental side effect** |
| --- | --- | --- |
| **Non-mint** | 1256 | 949 |
| **Mint** | 768 | 686 |

#### Dental professional survey

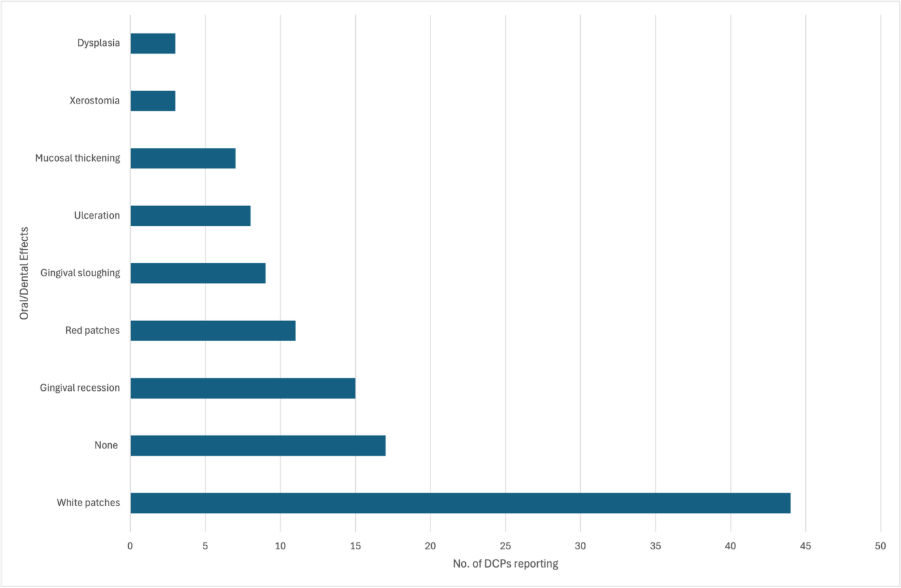

*Appendix Figure 2: Oral and dental effects of NPs reported by dental professionals.*

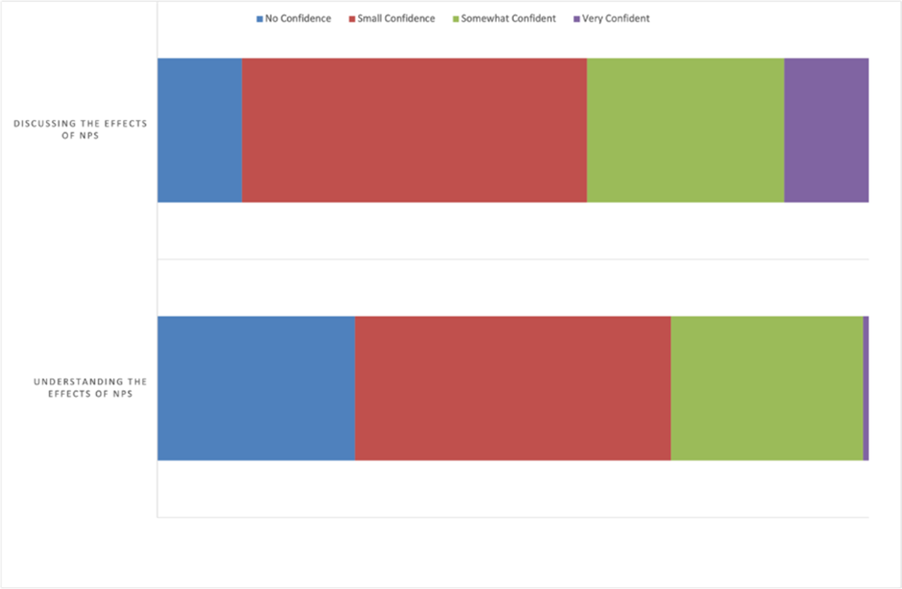

*Appendix Figure 3: Dental professionals’ confidence in understanding and discussing the effects of NPs with patients.*

*Appendix Table 5: Free text responses on advice/management:*

| **Management strategy** | **Example quotations** |
| --- | --- |
| Encouraging either cessation or reduction | *"To stop using them"*  *"Reduce frequency Reduce duration Reduce strength"*  *"Try to stop, reduce/wean. Seek help from NHS SSS or GP."*  *"I advise patient's of the unknown long term risk regarding use of nicotine pouches, and therefore would recommend for them to stop."*  *"I give cessation advice, but this may be tempered if they are using pouches in lieu of cigarettes as the risks of smoking are thought to be greater based on current evidence."* |
| Changing the site of NP pouch placement | *"Alternate areas of application, ideally stop using them"*  *"To rotate site they place the pouch in the mouth."*  *"Patients should be advised to rotate the nicotine pouch to different areas of the mouth, rather than placing it repeatedly in the same location."*  *"To try and stop using them, if they continue to use then move the pouches around and not hold in one place"* |
| Regular soft tissue monitoring | *"Monitor the site where they routinely place and report changes to the gum."*  *"If develops a white/red patch that does not heal after 2 weeks to contact the DC"*  *"To monitor the areas, to return for exam if any change in appearance or sensation lasting over 2 weeks."*  *"patients should be instructed to contact the dental centre promptly if they notice any changes, discomfort, irritation, or new sensations within the mouth."*  *"I advise these patients to check their mouths fortnightly for visible changes. If any white, red or speckled patches arise in the area of pouch use, to put the pouches somewhere else and check that the area resolves to normal. If it does not, to get this checked by a dentist as a matter of urgency."* |
| Discussion with patients about the potential risks of use and lack of current research | *"That it will probably adversely affect their mouth but we don’t know exactly how as they are pretty new"*  *"Not enough research has been done on pouches so we are still not aware of the extent of the negative effects that pouches could cause."*  *"They are quite a 'new' thing and we don't have much clinical data on them at the moment to guide us if they are dangerous or not."* |

*Appendix Table 6: Free text responses on carcinogenic question*

| **Carcinogenic** | **Example quotations** |
| --- | --- |
| Contents held in direct contact with soft tissues therefore potentially leading to cellular damage | *"Repeatedly damaging the mucosa, causing the risk of cell damage and possible uncontrollable cell division"*  *"Because over time they traumatise and reduce the blood supply to the tissue they are against."*  *"Constantly held against soft tissue. Chemicals - burning - aggravated tissue"*  *"Because of the risk of cellular change caused by contact with nicotine"* |
| Experience of current NP patients having biopsies showing dysplasia | *"Experience of mild dysplasia directly linked to use"*  *"I have seen multiple nicotine pouch related changes in the oral cavity and undertaken several biopsies which have shown varying degrees of keratosis/leukoplakia, with some returning cellular atypia and low grade dysplasia. This is concerning in the young demographic using these products"*  *"Encountered one patient (male, fit and well, early twenties) with dysplasia on biopsy of a gingival white patch at site of application."* |
| Confusion regarding NP contents | *"Due to the other products they contain, but I’m not 100% sure."*  *"due to small amounts of tobacco specific nitrosamines"*  *"Contain nitrosamines and formaldehyde"*  *"Tobacco"* |

*Appendix Table 7: Free text comments from dental professional’s survey*

| **Free text comment theme** | **Example quotations** |
| --- | --- |
| Rapid increase in use | *"Within the last year their use amongst my pt cohort - young, physically fit males, has increased exponentially"*  *"I work in a British Army base and it appears that the use of nicotine pouches in 18-40 year old men has significantly increased over the past c5 years especially with the site moving to be smoke-free."*  *"Appears to be increasingly trendy in younger generations compared with smoking and vaping. And used by those who wouldn’t typically use cigarettes."*  *"I have seen a lot of people (in work and socially) who use them who would never consider smoking cigarettes."* |
| Lack of long-term research and evidence | *"There isn't really much evidence yet."*  *"I have researched this quite extensively so I can give better information to patients but the evidence is so limiting."*  *"I think more mid-longterm research is required to understand the effects. I personally feel I do not have enough experience with these patients."*  *"There is insufficient evidence so far”* |
| Lack of regulations and public health concerns | *"The sector is completely unregulated”*  *"I am concerned with the rapid emergence of these products with little/no regulation, particularly in younger age groups."*  *"- I think the packaging making them look like sweets isn't good"*  *"needs to be health campaigns to deliver accurate information"*  *"The terminology/patient education needs to be pushed so that it's less 'cool'."* |
| Nicotine dependence and overuse | *"Patients report that they will use pouches as a way of stopping vaping or cigarettes but become more addicted to the pouches and it is harder to stop using them."*  *"One patient was using the equivalent of 300 cigarettes a day but previously only smoked 5 a day when he smoked when he tried to stop he felt very sick from withdrawals so has struggled to wean himself off again."*  *"Patients can increase their nicotine amount very easily without realising."*  *"They are very addictive and I found majority of patients use them now due to being able to do it at a desk."*  *"The main issue is overuse. I think per use they are less harmful than e-cig/cigarettes. However I find patients use the pouches far more often than e-cigs/cigarettes due to being able to use them in social situations far more easily."* |

### In-person assessment

*Appendix Table 8: Participant characteristics for in-person assessment.*

| **Characteristic** | **N (%)**  **(N=34)** |
| --- | --- |
| **Age in years, mean (sd; range))** | 27.5 (8.3; 18-44) |
| **Sex at birth, no. (%)** |  |
| Male | 28 (85%) |
| Female | 5 (15%) |
| **Ethnicity, no. (%)** |  |
| White | 26 (75%) |
| Asian or Asian British | 6 (17%) |
| Mixed or multiple ethnic groups | 1(3%) |
| Latino | 1(3%) |
| **Alcohol intake, in those that consume alcohol (n=23), mean units per week (sd; range)** | 13 (8.7; 1-30) |
| **Smoking status, no. (%)** |  |
| Current smoker | 10 (29%) |
| Former smoker | 19 (56%) |
| Never smoker | 5 (15%) |
| **Former smoker, how long (years) since stopping, mean (sd; range)** | 4.4 (3.0; 1-11) |
| **E-cigarette use, no. (%)** |  |
| Current user | 13 (38%) |
| Former user | 11 (32%) |
| Never user | 10 (29%) |

*Appendix Table 9: Oral health data from In-person assessments*

|  | | **Whole mouth** | **Pouch site** | **Non-pouch site** | **P value^a^** |
| --- | --- | --- | --- | --- | --- |
| **PPD (mm)** | Mean (SD) | 2.2 (0.4) | 2.0 (0.7) | 2.2 (0.4) | P=0.047 |
|  | Median (IQR) | 2.2 (1.9-2.5) | 2.0 (1.7-2.3) | 2.2 (1.9-2.5) |  |
|  | Range | (1.43,3.32) | (0.2,3.8) | (1.5,3.3) |  |
| **REC (mm)** | Mean (SD) | 0.2 (0.2) | 0.2 (0.3) | 0.1 (0.2) | P=0.991 |
|  | Median (IQR) | 0.1 (0.0-0.2) | 0 (0.0-0.3) | 0.1 (0.0-0.2) |  |
|  | Range | (0.0,1.0) | (0,1.3) | (0.0,1.0) |  |
| **Recession, % of sites** | Mean (SD) | 4.9 (8.5) | 3.0 (9.7) | 5.1 (9.2) | P=0.040 |
|  | Median (IQR) | 1.2 (0.0-7.7) | 0 (0-0) | 1.3 (0-8.4) |  |
|  | Range | (0,39.7) | (0,33.3) | (0,44.9) |  |
| **Any Recession** | % of participants | 65% |  | | |
| **BOP (%)** | Mean (SD) | 0.3 (0.2) | 0.2 (0.3) | 0.3 (0.2) | P=0.006 |
|  | Median (IQR) | 0.3 (0.2-0.5) | 0.0 (0.0-0.3) | 0.3 (0.2-0.5) |  |
|  | Range | (0.0,0.7) | (0.0,0.9) | (0.0,0.7) |  |
| **CAL (mm)** | Mean (SD) | 2.3 (0.5) | 2.2 (0.7) | 2.3 (0.6) | P=0.096 |
|  | Median (IQR) | 2.3 (2.0-2.6) | 2.0 (1.7-2.7) | 2.3 (2.0- 2.6) |  |
|  | Range | (1.4,4.3) | (0.7,3.8) | (1.5,4.4) |  |
| **% of sites with CAL ≥3mm** | Mean (SD) | 38.4 (20.4) | 26.3 (30.1) | 38.8 (20.5) | P=0.012 |
|  | Median (IQR) | 38.7 (21.9-50.9) | 23.5 (0-50) | 37.6 (22.5-51.2) |  |
|  | Range | (6,89.7) | (0,100) | (6.4,89.1) |  |
| **MGI** | Mean (SD) | 1.5 (0.7) | 1.4 (1.1) | 1.5 (0.7) | P=0.282 |
|  | Median (IQR) | 1.5 (0.9-2.1) | 1.0 (0.5-2.1) | 1.6 (0.9-2.1) |  |
|  | Range | (0.6,2.9) | (0.0,3.2) | (0.6,2.9) |  |
| **PI** | Mean (SD) | 0.5 (0.4) | 0.4 (0.6) | 0.6 (0.4) | P=0.004 |
|  | Median (IQR) | 0.5 (0.2-0.8) | 0.2 (0.0-0.5) | 0.5 (0.2-0.8) |  |
|  | Range | (0.0-1.9) | (0.0,2.7) | (0.0-1.8) |  |
| **CODS** | Mean (SD) | 0.1 (0.5) |  |  |  |
|  | Median (IQR) | 0.0 (0.0-0.0) |  |  |  |
|  | Range | (0,2) |  |  |  |

*a-Wilcoxon signed Rank Test*
