## Supplementary material for "Investigating the oral and dental effects of nicotine pouches: findings from a patient and professional survey with in-person assessment": Dental Professional Survey

### Dental Professionals Survey

The aim of this survey is to assess the impact of nicotine pouches on oral health. The survey is being ran by Newcastle University.

You can complete this survey if you are a dental professional.

The survey takes approximately 10 minutes to complete. Participants that complete the survey will have the option to enter a prize draw for a £100 gift voucher

The participant information sheet is available to view in full here.

This is the survey for dental professionals. If you are a member of the public who uses nicotine pouches there is a seperate survey for you here.

1. After reading the participant information statement do you consent to partake in the following survey?
   *

- [ ] Yes
- [ ] No

2. What does 2 + 2 equal
   *

[ ]

3. What is your current job role?
*

- ◯ General Dental Practitioner
- ◯ Specialist / StR
- ◯ Dental Therapist
- ◯ Dental Hygienist
- ◯ Dental Nurse
- ◯ Other (please specify)

4. Which specialist list are you on (or in training to go on)?
*

0/32,000 characters

5. What is the postcode of the practice/clinic/hospital you work at? (if multiple, please state the one you work at most frequently)
*

0/32,000 characters

6. Do you routinely ask patients whether they use nicotine pouches?
*

- ◯ Yes
- ◯ No

7. Is this question covered in your medical history form?
*

- ◯ Yes
- ◯ No

◯ Don't know

8. Please estimate the number of patients that you have seen that are self-reported users of nicotine pouches within the last year ?

*

◯ None
◯ 1-5
◯ 6-10
◯ 11-20
◯ 21-30
◯ +30

9. What advice would you give to patients that you encounter that use nicotine pouches?

*

0/32,000 characters

10. Please state, if any, changes that **<u>patients using nicotine pouches have reported</u>** to their oral hard or soft tissues

*

0/32,000 characters

11. Please state, if any, changes **<u>you have noticed</u>** to the oral hard or soft tissues of any patients that use nicotine pouches

*

0/32,000 characters

12. Compared to tobacco cigarettes do you believe that nicotine pouches are
*

- ◯ More harmful
- ◯ Less harmful
- ◯ About the same

13. Compared to e-cigarettes do you believe that nicotine pouches are
*

- ◯ More harmful
- ◯ Less harmful
- ◯ About the same

14. Do you believe that nicotine pouches are carcinogenic?
*

- ◯ Yes
- ◯ No
- ◯ Not sure

15. Please tell us why you think they might be carcinogenic?
*

0/32,000 characters

16. How confident do you feel in understanding the possible effects of oral nicotine pouches on oral and general health, based upon the current evidence?
*

- ◯ Not confident at all
- ◯ A small amount of confidence
- ◯ Somewhat confident

◯ Very confident

17. How confident do you feel in discussing the potential oral effects of nicotine pouch use with a patient?
*

◯ Not confident at all
◯ A small amount of confidence
◯ Somewhat confident
◯ Very confident

18. Is there anything else you would like to share with us about your experience of oral nicotine pouches?

0/32,000 characters

19. Thank you for taking the time to complete this survey. If you would like to be entered into the prize draw (£100 voucher) please provide your email address in the box below.

0/32,000 characters
