## Supplementary material for "Investigating the oral and dental effects of nicotine pouches: findings from a patient and professional survey with in-person assessment": Public Survey

### Oral nicotine pouch public survey

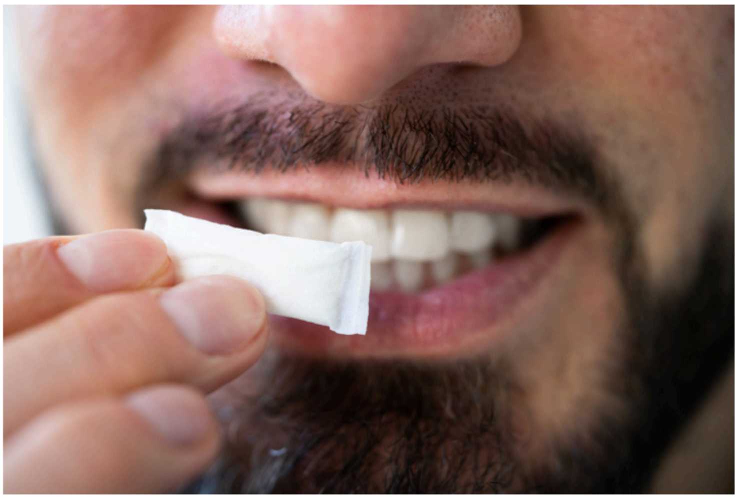

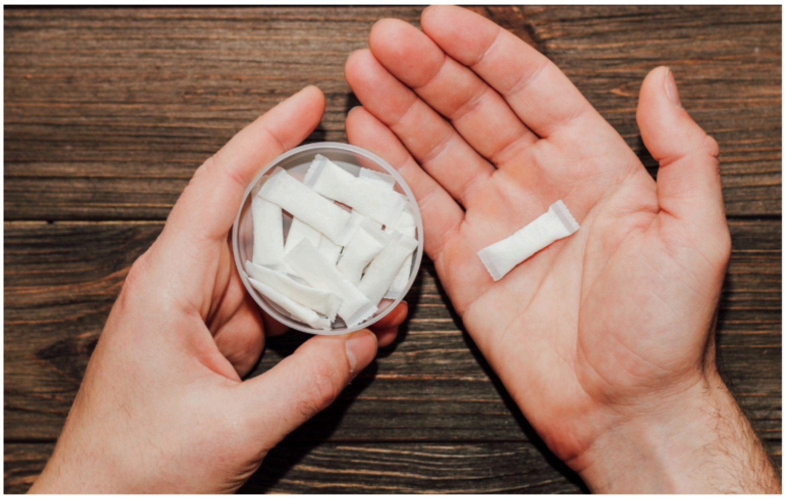

The aim of this public survey is to assess the impact of nicotine pouches on oral health. The survey is being ran by Newcastle University.

You can complete this survey if you regularly use oral nicotine pouches now or have done in the past.

The survey takes approximately 10-15 minutes to complete. Participants that complete the initial survey will have the option to enter a prize draw for a £100 gift voucher

The participant information sheet is available to view in full

This is the public survey, however if you are a dental professional you can access the nicotine pouches Dental professional survey on our website here.

After reading the participant information statement do you consent to partake in the following survey?

*

◯ Yes

○ No

What does 2+2 equal?
*

What is your age group?
*

- ◯ Under 16
- ◯ 16-18
- ◯ 19-22
- ◯ 23-25
- ◯ 26-30
- ◯ 31-40
- ◯ 41-50
- ◯ 51-60
- ◯ 61-70
- ◯ 71-80
- ◯ 81-90
- ◯ 91-100
- ◯ 101-110

This survey is looking at oral nicotine pouch use, which of the following product(s) do you use or have used previously? Select all that apply.
*

- ☐ Oral nicotine pouches (tobacco free, sometimes called 'white snus') e.g. ZYN, VELO, Nordic Spirit.
- ☐ Tobacco cigarettes
- ☐ E-cigarettes
- ☐ Snus (tobacco containing)
- ☐ Other

Do you currently use oral nicotine pouches (tobacco free) or have you stopped using this product? Select below
*

- ◯ I currently use oral nicotine pouches
- ◯ I previously used oral nicotine pouches but have now stopped

What is your postcode?
*

0/32,000 characters

Which of the following best describes your gender?
*

- Man
- Woman
- Non-binary
- Prefer not to say
- My gender is not listed here (please state)

What is your ethnicity?
*

- White - British
- White - Irish
- White - Gypsy or Irish Traveller
- Any other White background
- Mixed - White and Black Caribbean
- Mixed - White and Black African
- Mixed - White and Asian
- Asian/Asian British - Indian
- Asian/Asian British - Pakistani
- Asian/Asian British - Bangladeshi
- Asian/Asian British - Chinese
- Black/African/Caribbean/Black British - African
- Black/African/Caribbean/Black British - Caribbean
- Arab
- Any other ethnic group, (please specify)

What is your employment status?
*

- ◯ Employed
- ◯ Self-employed
- ◯ Retired
- ◯ In full time education
- ◯ Unemployed

What do you study?
*

0/32,000 characters

What is your occupation?
*

0/32,000 characters

Since you started using nicotine pouches have you noticed any side effects or changes to your mouth or teeth?
*

- ◯ Yes
- ◯ No

Which of the following have you noticed? Select all that apply
*

- ☐ Burning sensation
- ☐ Dry mouth
- ☐ Sore mouth
- ☐ Sore throat
- ☐ Mouth ulcers
- ☐ Gum recession/shrinkage
- ☐ White patches in mouth
- ☐ Other (please specify)

Since you started using nicotine pouches have you noticed any side effects or changes to your mouth or teeth?
*

◯ Yes
◯ No

Which of the following have you noticed? Select all that apply
*

☐ Burning sensation
☐ Dry mouth
☐ Sore mouth
☐ Sore throat
☐ Mouth ulcers
☐ Gum recession/shrinkage
☐ White patches in mouth
☐ Other (please specify)

In which areas of the diagram have you noticed this **burning sensation**? Select all that apply
*

- [ ] 1
- [ ] 2
- [ ] 3
- [ ] 4
- [ ] 5
- [ ] 6
- [ ] 7

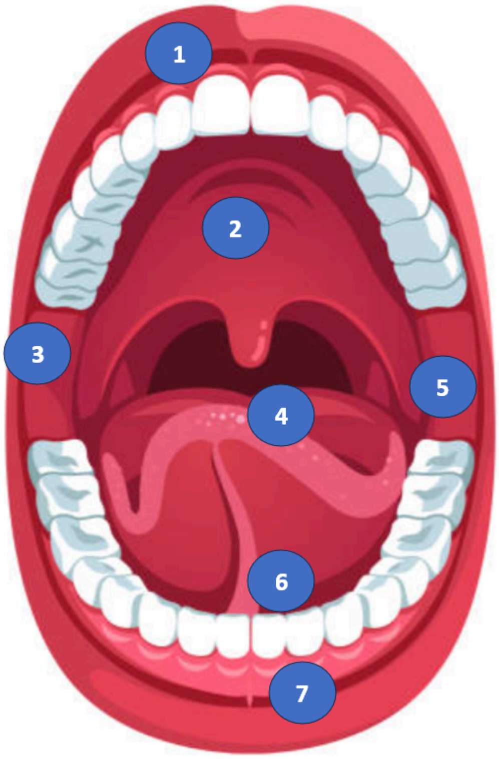


When do you notice this symptom in relation to using nicotine pouches?
*

- ◯ During use
- ◯ Immediately after use
- ◯ For an extended time after use
- ◯ Permanently present

○ Other (please specify)

On a scale of 1 to 10 how much does this symptom bother you? (1 being least bothered and 10 being most bothered)
*

○ 1
○ 2
○ 3
○ 4
○ 5
○ 6
○ 7
○ 8
○ 9
○ 10

In which areas of the diagram did you notice this **burning sensation**? Select all that apply
*

- [ ] 1
- [ ] 2
- [ ] 3
- [ ] 4
- [ ] 5
- [ ] 6
- [ ] 7

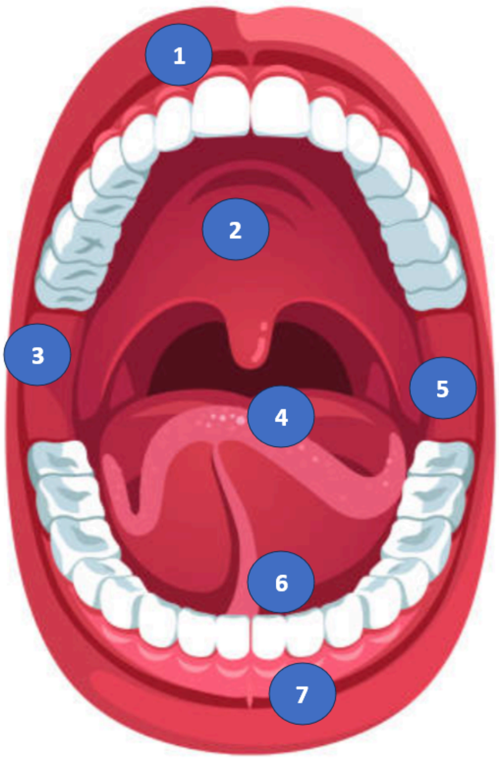


When did you you notice this symptom in relation to using nicotine pouches?
*

- ◯ During use
- ◯ Immediately after use
- ◯ For an extended time after use
- ◯ Permanently present
- ◯ Other (please specify)

On a scale of 1 to 10 how much did this symptom bother you? (1 being least bothered and 10 being most bothered)
*

- 1
- 2
- 3
- 4
- 5
- 6
- 7
- 8
- 9
- 10

When do you notice your **<u>mouth is dry</u>** in relation to using nicotine pouches?
*

- ◯ During use
- ◯ Immediately after use
- ◯ For an extended time after use
- ◯ Permanently present
- ◯ Other (please specify)

On a scale of 1 to 10 how much does this symptom bother you? (1 being least bothered and 10 being most bothered)
*

- ◯ 1
- ◯ 2
- ◯ 3
- ◯ 4
- ◯ 5
- ◯ 6
- ◯ 7
- ◯ 8
- ◯ 9
- ◯ 10

When did you notice your **<u>mouth was dry</u>** in relation to using nicotine pouches?
*

- ◯ During use
- ◯ Immediately after use
- ◯ For an extended time after use
- ◯ Permanently present
- ◯ Other (please specify)

On a scale of 1 to 10 how much did this symptom bother you? (1 being least bothered and 10 being most bothered)
*

- ◯ 1
- ◯ 2
- ◯ 3
- ◯ 4
- ◯ 5
- ◯ 6
- ◯ 7
- ◯ 8
- ◯ 9
- ◯ 10

In which areas of the diagram have you felt your **mouth is sore**? Select all that apply
*

- ☐ 1
- ☐ 2
- ☐ 3
- ☐ 4
- ☐ 5
- ☐ 6
- ☐ 7

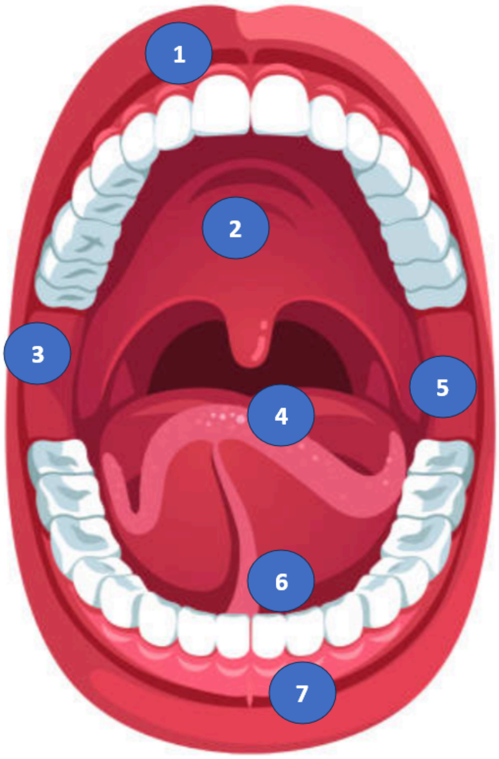


When do you notice this symptom in relation to using nicotine pouches?
*

- ◯ During use
- ◯ Immediately after use
- ◯ For an extended time after use
- ◯ Permanently present
- ◯ Other (please specify)

On a scale of 1 to 10 how much does this symptom bother you? (1 being least bothered and 10 being most bothered)
*

- ◯ 1
- ◯ 2
- ◯ 3
- ◯ 4
- ◯ 5
- ◯ 6
- ◯ 7
- ◯ 8
- ◯ 9
- ◯ 10

In which areas of the diagram did you feel your **mouth was sore**? Select all that apply
*

- [ ] 1
- [ ] 2
- [ ] 3
- [ ] 4
- [ ] 5
- [ ] 6
- [ ] 7

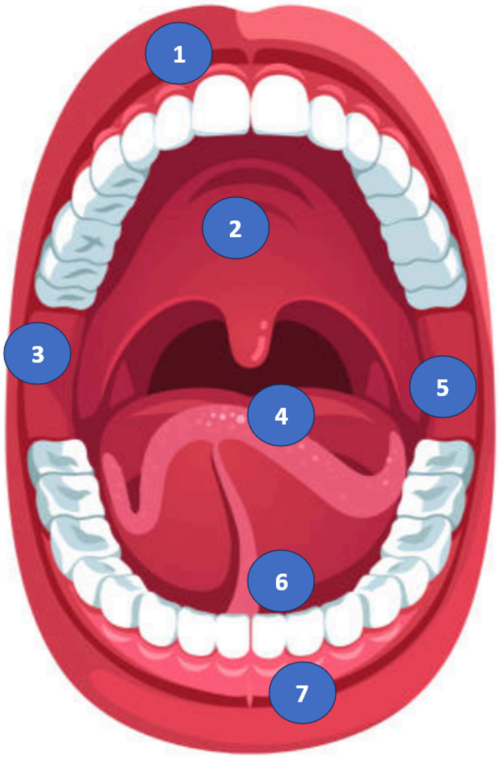


When did you notice this symptom in relation to using nicotine pouches?
*

- ◯ During use
- ◯ Immediately after use
- ◯ For an extended time after use
- ◯ Permanently present
- ◯ Other (please specify)

On a scale of 1 to 10 how much did this symptom bother you? (1 being least bothered and 10 being most bothered)
*

- ◯ 1
- ◯ 2
- ◯ 3
- ◯ 4
- ◯ 5
- ◯ 6
- ◯ 7
- ◯ 8
- ◯ 9
- ◯ 10

When do you notice that your **<u>throat is sore</u>** in relation to using nicotine pouches?
*

- ◯ During use
- ◯ Immediately after use
- ◯ For an extended time after use
- ◯ Permanently present
- ◯ Other (please specify)

On a scale of 1 to 10 how much does this symptom bother you? (1 being least bothered and 10 being most bothered)
*

- ◯ 1
- ◯ 2
- ◯ 3
- ◯ 4
- ◯ 5
- ◯ 6
- ◯ 7
- ◯ 8
- ◯ 9
- ◯ 10

When did you notice that your **throat was sore** in relation to using nicotine pouches?
*

- ◯ During use
- ◯ Immediately after use
- ◯ For an extended time after use
- ◯ Permanently present
- ◯ Other (please specify)

On a scale of 1 to 10 how much did this symptom bother you? (1 being least bothered and 10 being most bothered)
*

- ◯ 1
- ◯ 2
- ◯ 3
- ◯ 4
- ◯ 5
- ◯ 6
- ◯ 7
- ◯ 8
- ◯ 9
- ◯ 10

In which areas of the diagram have you noticed **mouth ulcers**? Select all that apply
*

- ☐ 1
- ☐ 2
- ☐ 3
- ☐ 4
- ☐ 5
- ☐ 6
- ☐ 7

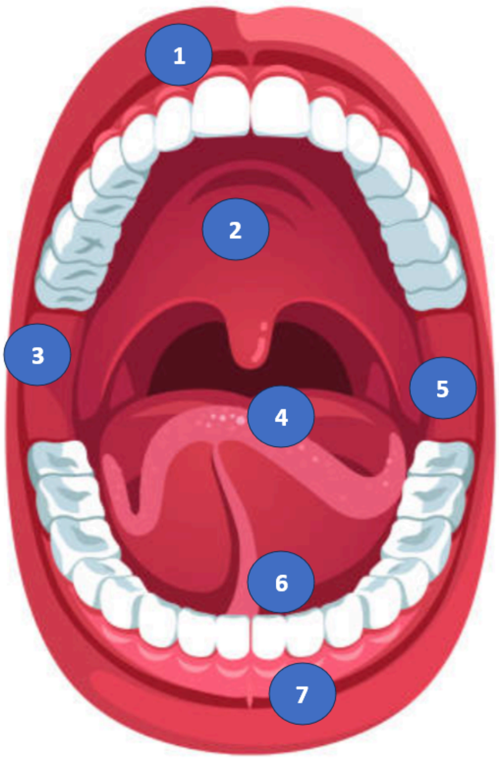


When do you notice this symptom in relation to using nicotine pouches?
*

- ◯ During use
- ◯ Immediately after use
- ◯ For an extended time after use
- ◯ Permanently present
- ◯ Other (please specify)

On a scale of 1 to 10 how much does this symptom bother you? (1 being least bothered and 10 being most bothered)
*

- 1
- 2
- 3
- 4
- 5
- 6
- 7
- 8
- 9
- 10

In which areas of the diagram did you notice **mouth ulcers**? Select all that apply

*

- [ ] 1
- [ ] 2
- [ ] 3
- [ ] 4
- [ ] 5
- [ ] 6
- [ ] 7

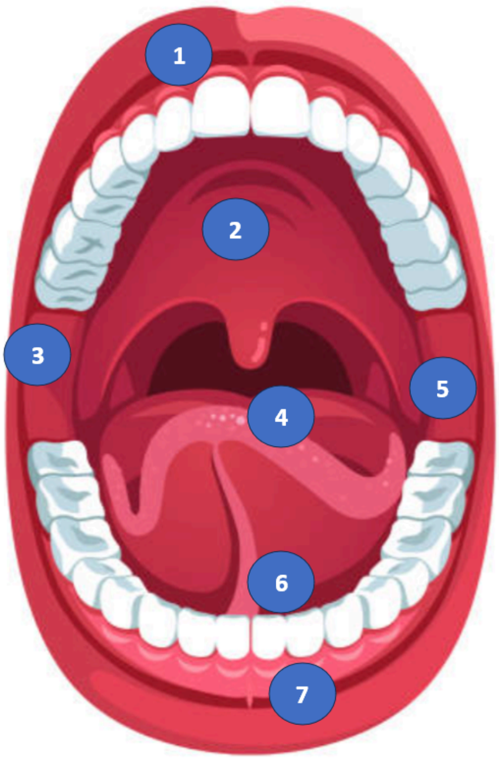


When did you notice this symptom in relation to using nicotine pouches?

*

- ◯ During use
- ◯ Immediately after use
- ◯ For an extended time after use
- ◯ Permanently present
- ◯ Other (please specify)

On a scale of 1 to 10 how much did this symptom bother you? (1 being least bothered and 10 being most bothered)
*

- ◯ 1
- ◯ 2
- ◯ 3
- ◯ 4
- ◯ 5
- ◯ 6
- ◯ 7
- ◯ 8
- ◯ 9
- ◯ 10

In which areas of the diagram have you noticed **<u>gum recession</u>** or shrinkage? Select all that apply

*

- [ ] 1
- [ ] 2
- [ ] 3
- [ ] 4
- [ ] 5
- [ ] 6
- [ ] 7

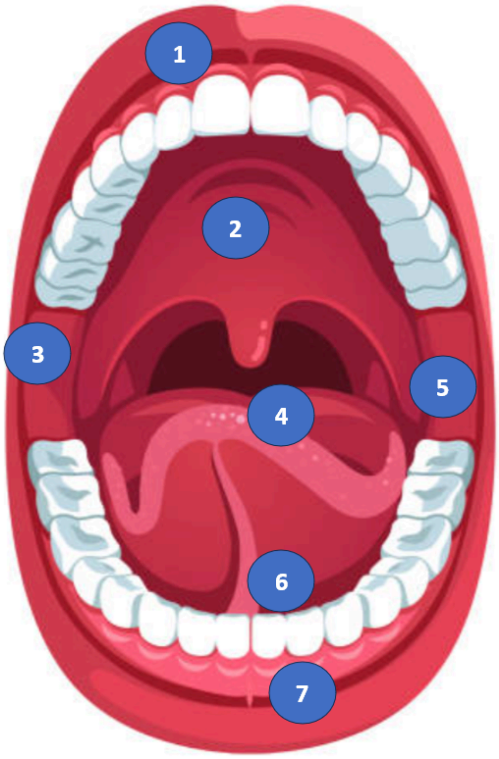


When do you notice this symptom in relation to using nicotine pouches?

*

- ○ During use
- ○ Immediately after use
- ○ For an extended time after use
- ○ Permanently present

- [ ] Other (please specify)

On a scale of 1 to 10 how much does this symptom bother you? (1 being least bothered and 10 being most bothered)
*

- [ ] 1
- [ ] 2
- [ ] 3
- [ ] 4
- [ ] 5
- [ ] 6
- [ ] 7
- [ ] 8
- [ ] 9
- [ ] 10

In which areas of the diagram have you **gum recession** or shrinkage? Select all that apply
*

- [ ] 1
- [ ] 2
- [ ] 3
- [ ] 4
- [ ] 5
- [ ] 6
- [ ] 7

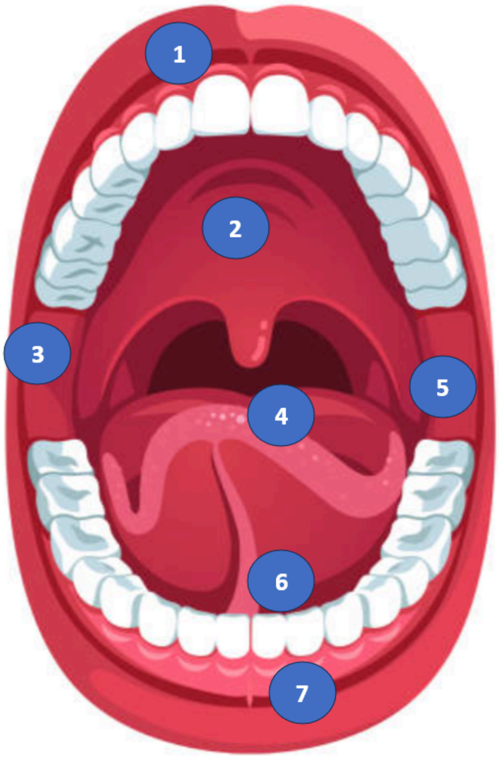


When did you notice this symptom in relation to using nicotine pouches?
*

- ○ During use
- ○ Immediately after use
- ○ For an extended time after use
- ○ Permanently present
- ○ Other (please specify)

On a scale of 1 to 10 how much does this symptom bother you? (1 being least bothered and 10 being most bothered)
*

- ◯ 1
- ◯ 2
- ◯ 3
- ◯ 4
- ◯ 5
- ◯ 6
- ◯ 7
- ◯ 8
- ◯ 9
- ◯ 10

In which areas of the diagram have you noticed a **white patch**? Select all that apply
*

- ☐ 1
- ☐ 2
- ☐ 3
- ☐ 4
- ☐ 5
- ☐ 6
- ☐ 7

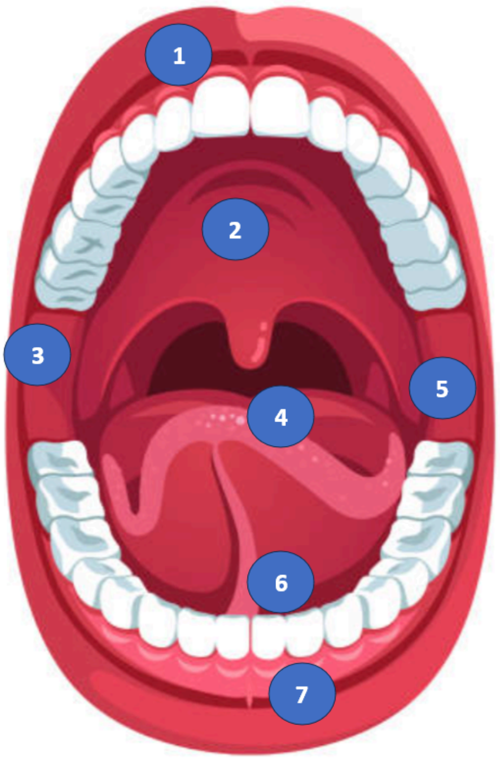


When do you notice this symptom in relation to using nicotine pouches?
*

- ◯ During use
- ◯ Immediately after use
- ◯ For an extended time after use
- ◯ Permanently present
- ◯ Other (please specify)

On a scale of 1 to 10 how much does this symptom bother you? (1 being least bothered and 10 being most bothered)
*

- ◯ 1
- ◯ 2
- ◯ 3
- ◯ 4
- ◯ 5
- ◯ 6
- ◯ 7
- ◯ 8
- ◯ 9
- ◯ 10

In which areas of the diagram have you noticed a **white patch**? Select all that apply

*

- [ ] 1
- [ ] 2
- [ ] 3
- [ ] 4
- [ ] 5
- [ ] 6
- [ ] 7

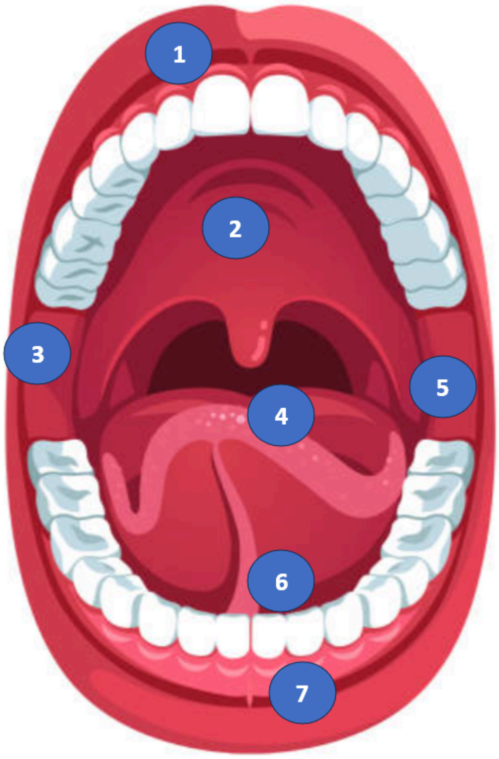


When did you notice this symptom in relation to using nicotine pouches?

*

- ◯ During use
- ◯ Immediately after use
- ◯ For an extended time after use
- ◯ Permanently present
- ◯ Other (please specify)

On a scale of 1 to 10 how much does this symptom bother you? (1 being least bothered and 10 being most bothered)
*

- 1
- 2
- 3
- 4
- 5
- 6
- 7
- 8
- 9
- 10

In which areas of the diagram have you noticed this symptom? Select all that apply
*

- [ ] 1
- [ ] 2
- [ ] 3
- [ ] 4
- [ ] 5
- [ ] 6
- [ ] 7

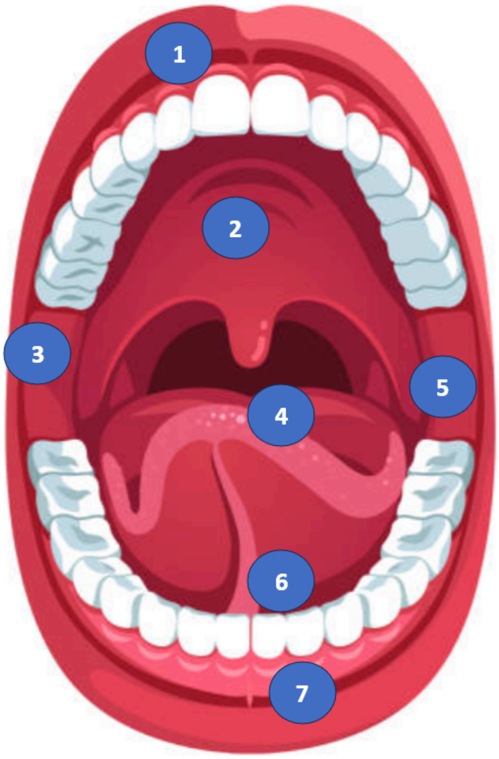


When do you notice this symptom in relation to using nicotine pouches?
*

- ○ During use
- ○ Immediately after use
- ○ For an extended time after use
- ○ Permanently present
- ○ Other (please specify)

On a scale of 1 to 10 how much does this symptom bother you? (1 being least bothered and 10 being most bothered)
*

- 1
- 2
- 3
- 4
- 5
- 6
- 7
- 8
- 9
- 10

Would you be willing to take a photograph of any lesion(s) you have experienced in your mouth on your smart phone or other device and upload it to this survey?
*

- ◯ Yes
- ◯ No

Please upload the photo of the lesion you have experienced in your mouth below
*

Select file

Would you like to upload another photo of any lesions you have experienced?
*

- ◯ Yes
- ◯ No

Please upload a second photo of any lesions you have experienced in your mouth below
*

Select file

Would you like to upload a third photo of any lesions you have experienced?
*

- ◯ Yes
- ◯ No

Please upload a third photo of any lesions you have experienced in your mouth below
*

Select file

In which areas of the diagram have you noticed this symptom? Select all that apply
*

- ☐ 1
- ☐ 2
- ☐ 3
- ☐ 4
- ☐ 5
- ☐ 6
- ☐ 7

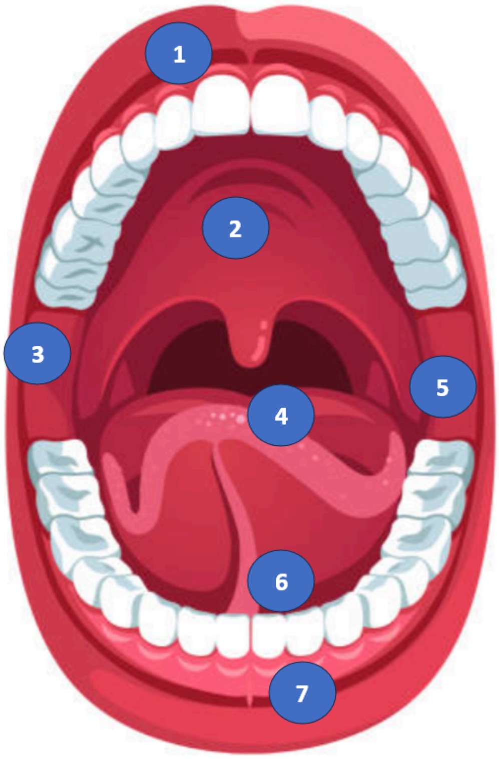


When did you notice this symptom in relation to using nicotine pouches?
*

- ◯ During use
- ◯ Immediately after use
- ◯ For an extended time after use
- ◯ Permanently present
- ◯ Other (please specify)

On a scale of 1 to 10 how much did this symptom bother you?
*

- 1
- 2
- 3
- 4
- 5
- 6
- 7
- 8
- 9
- 10

Would you be willing to take a photograph of any lesion(s) you have experienced in your mouth on your smart phone or other device and upload them?
*

- ◯ Yes
- ◯ No

Please upload the photo of any lesion you have experienced in your mouth below
*

Select file

Would you like to upload another photo of any lesions you have experienced?
*

- ◯ Yes
- ◯ No

Please upload a second photo of any lesions you have experienced in your mouth below
*

Select file

Would you like to upload a third photo of any lesions you have experienced?
*

- ◯ Yes
- ◯ No

Please upload a third photo of any lesions you have experienced in your mouth below
*

Select file

How long have you been using oral nicotine pouches?
*

- ◯ Less than one month
- ◯ One month to six months
- ◯ Six months to one year
- ◯ One to two years
- ◯ Two to five years
- ◯ More than five years

How many fresh nicotine pouches do you typically use per day?
*

- ◯ 1
- ◯ 2 to 5
- ◯ 6 to 10
- ◯ 11 to 20
- ◯ More than 20

What time of day do you tend to use nicotine pouches? Select all that apply?
*

- ☐ Morning
- ☐ Mid morning
- ☐ Lunchtime
- ☐ Afternoon
- ☐ Early evening
- ☐ Night time

How many hours per day do you have a nicotine pouch in your mouth?
*

- ◯ Less than 1 hour
- ◯ 1-2 hours
- ◯ 3-4 hours
- ◯ 5-6 hours
- ◯ 7-8 hours

- ◯ 9-10 hours
- ◯ 11-12 hours
- ◯ 13-14 hours
- ◯ 15-16 hours
- ◯ More than 16 hours

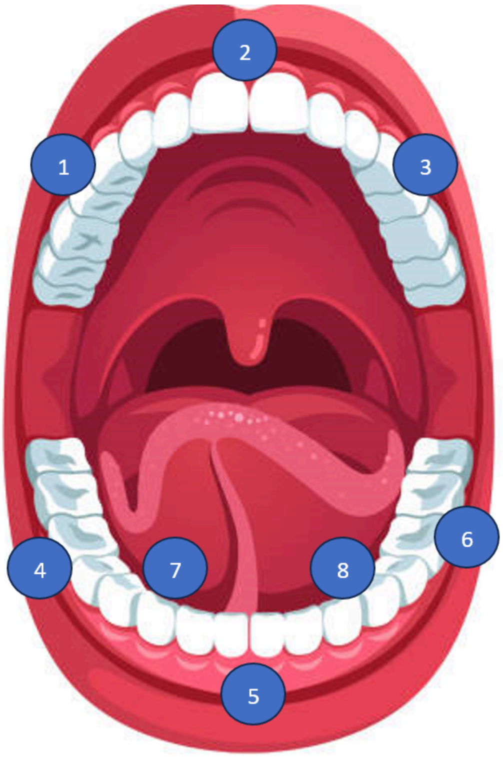


Where do you typically hold the nicotine pouch in your mouth on the diagram? Select all that apply
*

- ☐ 1
- ☐ 2
- ☐ 3
- ☐ 4
- ☐ 5
- ☐ 6
- ☐ 7
- ☐ 8

What brand of nicotine pouches do you use most often?

0/32,000 characters

What flavour of nicotine pouches do you use most often?

0/32,000 characters

What strength of nicotine pouches do you use most often?
*

0/32,000 characters

Where do you get your nicotine pouches? Select all that apply
*

- [ ] Shops
- [ ] Online
- [ ] Social media e.g. Instagram, Tiktok shop
- [ ] Whatsapp
- [ ] Family
- [ ] Friends
- [ ] Other (please specify)

For how long did you use oral nicotine pouches?
*

- ◯ Less than one month
- ◯ One month to six months
- ◯ Six months to one year
- ◯ One to two years
- ◯ Two to five years
- ◯ More than five years

For how long have you stopped use oral nicotine pouches?
*

- ◯ Less than one month
- ◯ One month to six months
- ◯ Six months to one year
- ◯ One to two years
- ◯ Two to five years
- ◯ More than five years

How many fresh nicotine pouches did you typically use per day?
*

- ◯ 1
- ◯ 2 to 5
- ◯ 6 to 10
- ◯ 11 to 20
- ◯ More than 20

What time of day did you tend to use nicotine pouches? Select all that apply?
*

- ☐ Morning
- ☐ Mid morning
- ☐ Lunchtime
- ☐ Afternoon
- ☐ Early evening

- [ ] Night time

How many hours per day did you typically have a nicotine pouch in your mouth?
*

- ( ) Less than 1 hour
- ( ) 1-2 hours
- ( ) 3-4 hours
- ( ) 5-6 hours
- ( ) 7-8 hours
- ( ) 9-10 hours
- ( ) 11-12 hours
- ( ) 13-14 hours
- ( ) 15-16 hours
- ( ) More than 16 hours

What brand of nicotine pouches do you use most often?

0/32,000 characters

What flavour of nicotine pouches did you use most often?

0/32,000 characters

What strength of nicotine pouches did you use most often?
*

0/32,000 characters

Where did you get your nicotine pouches? Select all that apply
*

- [ ] Shops
- [ ] Online

- [ ] Social media e.g. Instagram, Tiktok shop
- [ ] Whatsapp
- [ ] Family
- [ ] Friends
- [ ] Other (please specify)

Do the nicotine pouches that you use contain tobacco?
*

- ◯ Yes
- ◯ No
- ◯ Not sure

To what extent do you agree or disagree with the following statements
*

I understand what nicotine pouches contain

- ◯ Strongly disagree
- ◯ Disagree
- ◯ Neutral
- ◯ Agree
- ◯ Strongly agree

I understand the benefits of using nicotine pouches

- ◯ Strongly disagree
- ◯ Disagree
- ◯ Neutral
- ◯ Agree
- ◯ Strongly agree

I understand the risks and/or side effects of using nicotine pouches

- ◯ Strongly disagree
- ◯ Disagree
- ◯ Neutral
- ◯ Agree
- ◯ Strongly agree

I want to know more about what nicotine pouches contain

- ◯ Strongly disagree
- ◯ Disagree
- ◯ Neutral
- ◯ Agree
- ◯ Strongly agree

I want to know more about the benefits of using nicotine pouches

- ◯ Strongly disagree
- ◯ Disagree
- ◯ Neutral
- ◯ Agree
- ◯ Strongly agree

I want to know more about the risks/side effects of using nicotine pouches

- ◯ Strongly disagree
- ◯ Disagree
- ◯ Neutral
- ◯ Agree
- ◯ Strongly agree

In terms of what you currently know about nicotine pouches, where have you learnt this? Select all that apply
*

- ☐ Online
- ☐ Social media
- ☐ Work
- ☐ School
- ☐ Friends or family
- ☐ Doctor or other health care professional
- ☐ Other (please specify)

Do the nicotine pouches that you used contain tobacco?
*

- Yes
- No
- Not sure

To what extent do you agree or disagree with the following statements
*

I understand what nicotine pouches contain

- Strongly disagree
- Disagree
- Neutral
- Agree
- Strongly agree

I understand the benefits of using nicotine pouches

- Strongly disagree
- Disagree
- Neutral
- Agree
- Strongly agree

I understand the risks and/or side effects of using nicotine pouches

- Strongly disagree
- Disagree
- Neutral
- Agree
- Strongly agree

I want to know more about what nicotine pouches contain

- Strongly disagree
- Disagree
- Neutral
- Agree
- Strongly agree

I want to know more about the benefits of using nicotine pouches

- ◯ Strongly disagree
- ◯ Disagree
- ◯ Neutral
- ◯ Agree
- ◯ Strongly agree

I want to know more about the risks/side effects of using them

- ◯ Strongly disagree
- ◯ Disagree
- ◯ Neutral
- ◯ Agree
- ◯ Strongly agree

In terms of what you currently know about nicotine pouches, where have you learnt this? Select all that apply
*

- ☐ Online
- ☐ Social media
- ☐ Work
- ☐ School
- ☐ Friends or family
- ☐ Doctor or other health care professional
- ☐ Other (please specify)

Have you noticed any of the following side effects when using oral nicotine pouches?
*

- [ ] Anxiety
- [ ] Vomiting
- [ ] Headaches
- [ ] Difficulty sleeping
- [ ] Tight chest
- [ ] I have not experienced any side effects
- [ ] Other (please specify)

Have you consulted a healthcare professional about these side effects?
*

- ( ) Yes
- ( ) No

Who have you seen?
*

- [ ] GP
- [ ] Pharmacist
- [ ] Dentist
- [ ] Other (please specify)

We are inviting some participants to the dental hospital in Newcastle for a dental check-up to assess the effects of using nicotine pouches on your mouth. Participants will receive a £50 voucher. Are you interested in being contacted about this?
*

- ○ Yes
- ○ No

Please provide your **email address:**
*

0/32,000 characters

Please provide your **contact number:**
*

0/32,000 characters

Please provide your **address:**

0/32,000 characters

#### Dental Monitoring/Photographs

We are also inviting some participants to receive a home monitoring kit (this includes a holder that clips to your phone and allows you to take photos inside your mouth which we can access). Should you be selected there will be no charge to you. You will receive a £20 voucher once you have completed the photos/scan. You will need to download an app on your smartphone.

Would you be interested in receiving a home monitoring kit through the post to provide some photographs of your mouth?
*

- ○ Yes
- ○ No

If you are interested in receiving this home monitoring kit please provide your postal address:
*

0/32,000 characters

Did you hear about this survey through advertising on Facebook or Instagram?
*

- ◯ Yes
- ◯ No

Thank you for taking the time to complete this survey. If you would like to be entered into the prize draw (£100 voucher) please provide your email address in the box below.

0/32,000 characters
